# Neural stimulation and recording performance in human somatosensory cortex over 1500 days

**DOI:** 10.1101/2020.01.21.20018341

**Authors:** Christopher L. Hughes, Sharlene N. Flesher, Jeffrey M. Weiss, John E. Downey, Jennifer L. Collinger, Robert A. Gaunt

## Abstract

**Objective:** Intracortical microstimulation (ICMS) in somatosensory cortex can restore sensation to people who have lost it due to spinal cord injury or other conditions. One potential challenge for chronic ICMS is whether neural recording and stimulation can remain stable over many years. This is particularly relevant since the recording quality of implanted microelectrode arrays frequently experience degradation over time and stimulation safety has been considered a potential barrier to the clinical use of ICMS. Our objective is to evaluate stability of recordings on intracortical stimulated and non-stimulated electrodes in a human participant across a long period of implantation. Additionally, we would like to assess the ability to evoke sensations with ICMS over time.

**Approach:** In a study investigating intracortical implants for a bidirectional brain-computer interface, we implanted microelectrode arrays with sputtered iridium oxide tips in the somatosensory cortex of a human participant with a cervical spinal cord injury. We regularly stimulated through electrodes on these microelectrode arrays to evoke tactile sensations on the hand. Here, we quantify the stability of these electrodes in comparison to non-stimulated electrodes implanted in motor cortex over 1500 days in two ways: recorded signal quality and electrode impedances. Additionally, we quantify the perceptual stability of ICMS-evoked sensations with detection thresholds.

**Main results:** We found that recording quality, as assessed by the number of electrodes with high-amplitude waveform recordings (> 100 µV), peak-to-peak voltage, noise, and signal-to-noise ratio, generally decreased over time on stimulated and non-stimulated electrodes. However, stimulated electrodes were much more likely to continue to record high-amplitude signals than non-stimulated electrodes. Interestingly, the detection thresholds for stimulus-evoked tactile sensations decreased over time from a median of 31.5 μA at Day 100 to 10.4 μA at Day 1500, with the most substantial changes occurring between Day 100 and Day 500.

**Significance:** These results provide evidence that ICMS in human somatosensory cortex can be provided over long periods of time without deleterious effects on recording or stimulation capabilities. In fact, psychophysical sensitivity to stimulation improves over time and stimulation itself may promote more robust long-term neural recordings.

## 1. Introduction

Brain-computer interfaces (BCIs) can be used to restore functional motor control by connecting the brain to an effector or assistive device and bypassing the spinal cord and limbs. While methods such as electroencephalography and electrocorticography can be used as the basis of a BCI [1], [2], penetrating intracortical electrode arrays currently allow for the highest resolution recordings due to the proximity of the tips to the neural population from which they are recording. Indeed, in recent years intracortical implants in motor cortex have been used for high-performance BCI control [3]–[7]. Additionally, electrical microstimulation through intracortical implants in somatosensory cortex has been used to generate tactile percepts in people with chronic spinal cord injury [8], [9]. As these devices must be surgically implanted, microelectrode arrays need to function for long periods of time to be clinically practical.

The stability of microelectrode arrays in motor cortex has been well studied in non-human primates [10]–[12]. Additionally, there are several reports on signal quality and stability for electrodes implanted in human motor cortex [5], [13]–[15]. Although intersubject variability is high, signals can be reliably recorded for up to 3-5 years when devices do not fail, although the quality of these recordings deteriorates over time. However, whether electrodes primarily used for intracortical microstimulation (ICMS) can record and stimulate effectively over clinically relevant timelines is uncertain, especially given that stimulation itself may have detrimental effects on the electrodes and the surrounding neural tissue [16]– [19].

This concern is particularly relevant for people, since implantation requires a craniotomy performed under general anesthesia. In order to study the sensations evoked by ICMS in human somatosensory cortex, we implanted intracortical microelectrode arrays into the somatosensory cortex of a participant with C5/C6 spinal cord injury. With these devices, we have demonstrated that ICMS can elicit tactile percepts localized to the hand on the majority of implanted electrodes [8]. Prior to this clinical study, a series of experiments in non-human primates were performed to establish safe stimulation limits using microelectrodes arrays. These studies demonstrated that frequent microstimulation over six months did not appear to cause more damage to the cortex than what could be expected from implanting the devices themselves, that there were minimal effects on the characteristics of the electrode-tissue interface, and that that there were no detectable behavioral changes in the animals’ abilities to perform dexterous behaviors that require somatosensory feedback [20], [21]. Other studies have also demonstrated that signal can be reliably recorded and sensations can be reliably evoked over long periods of time in animals when using stimulation amplitudes up to 300 μA [18], [21]–[23].

Despite the precautions taken in this study to use parameters that have proven effective in animals, it is necessary to demonstrate that these parameters are safe and effective in humans. Here, we present results on the stability of Utah microelectrode arrays used for ICMS in the somatosensory cortex of a human participant over 1500 days using these safety standards derived from non-human primate studies.

## 2. Materials and Methods

Data for this study were collected for the first 1500 days after implantation. Typically, experiments were conducted three days a week. Experimental sessions were typically four-hours long, although stimulation was not used in every session or for all four hours. Across the 1500 implant days, experiment sessions were performed on 510 days, 378 of which involved stimulation.

### 2.1 Participant and Implants

This study was conducted under an Investigational Device Exemption from the U.S. Food and Drug administration, approved by the Institutional Review Boards at the University of Pittsburgh (Pittsburgh, PA) and the Space and Naval Warfare Systems Center Pacific (San Diego, CA), and registered at ClinicalTrials.gov (NCT0189-4802). Informed consent was obtained before any study procedures were conducted. A single subject was implanted with two microelectrode arrays (Blackrock Microsystems, Salt Lake City, UT) in somatosensory cortex [8]. Each electrode array consisted of 32 wired electrodes distributed throughout a 6×10 grid. All 60 electrodes were not wired due to technical constraints related to the total available number of electrical contacts. Electrode tips were coated with a sputtered iridium oxide film (SIROF) [8]. The participant also had two microelectrode arrays implanted in motor cortex. Each electrode array consisted of 88 wired electrodes distributed throughout a 10×10 grid. Electrode tips in motor cortex were coated with platinum. Additional details on these implants can be found in other studies focusing on the motor and sensory performance of these devices [4], [24]. It should be noted that there are differences in the cellular composition of motor and somatosensory cortex, most notably the presence of large Betz cells in motor cortex which can produce larger signal [25]. Differences in recordings could then be affected by cortical differences regardless of stimulation.

### 2.2 Stimulation protocol

Stimulation was delivered using a CereStim C96 multichannel microstimulation system (Blackrock Microsystems, Salt Lake City, UT). Pulse trains consisted of cathodal phase first, current-controlled, charge-balanced pulses delivered at frequencies from 20-300 Hz and at amplitudes from 2-100 μA. The cathodal phase was 200 μs long, the anodal phase was 400 μs long, and the anodal phase was set to half the amplitude of the cathodal phase. The phases were separated by a 100 μs interphase period. At the beginning of each test session involving stimulation, we sequentially stimulated each electrode first at 10 μA and 100 Hz for 0.5 s and then at 20 μA and 100 Hz for 0.5 s. During these trials, the interphase voltage on each electrode was measured at the end of the interphase period, immediately before the anodal phase [26]. If an electrode’s measured interphase voltage was below -1.5 V, the electrode was disabled for the day. This step was performed to minimize stimulation on electrodes that might potentially experience high voltages, which could result in irreversible damage. Typically, 2 to 8 electrodes were removed from the test set each day. The electrodes that were removed tended to change from day to day suggesting that the high interphase voltage was due to poor contact between the percutaneous connector and the stimulation cable. On any given day, tactile sensations localized to the hand were elicited on 50-60 of the 64 electrodes with stimulation [8]. All electrodes, except those exhibiting high interphase voltages, were stimulated at suprathreshold amplitudes, typically 60 μA, at least once a month during a monthly survey of elicited tactile sensations. Some electrodes were used more frequently for other tasks involving stimulation.

We conducted experiments exclusively at the participant’s home from Day 364 to 498 for reasons unrelated to the study. Experiments conducted in the home used a different set of hardware and were subject to different environmental noise characteristics than the laboratory. Stimulation experiments were not conducted during this time because our regulatory approvals dictated that stimulation may only be performed in the lab. However, neural recordings were made from electrodes implanted in somatosensory cortex during this time and those data are included in this manuscript. After implant Day 498, stimulation experiments continued both at home and in the laboratory after regulatory modifications had been approved.

### 2.3 Stability metrics

To quantify the stability of stimulated electrodes in somatosensory cortex, we measured recorded signal quality, electrode impedances, and stimulation detection thresholds over time. Signal quality and impedance were compared between stimulated electrodes and non-stimulated electrodes. Recording quality was evaluated using four metrics: (1) the number of high-amplitude recordings, (2) peak-to-peak voltage, (3) noise, and (4) signal-to-noise ratio. Ideally, a clinical device would be able to record many neurons with large peak-to-peak voltages, have low noise levels, and therefore, a high signal-to-noise ratio for the duration of the implant.

Another key metric of electrode stability, particularly for stimulated electrodes, is the electrode impedance. Increases in the impedance could result in higher required voltages at the electrode-tissue interface for constant-current stimulation pulses, resulting in potentially irreversible reduction and oxidation reactions [26]. Therefore, it is necessary for electrode impedances to remain stable over the length of the implant.

Finally, the ability to elicit sensations with ICMS must necessarily be maintained. Detection thresholds are a measure of how well the participant can detect electrical stimulation. Changes in this parameter reflect the amount of charge that must be used to induce tactile percepts, and there is an upper limit to the amount of charge that can be safely applied [19]. Therefore, detection thresholds should not systematically increase over the length of implantation. Increasing thresholds may then result in an inability to evoke tactile percepts with parameters that have been shown to be safe.

### 2.4 Neural recordings

At the beginning of each session, we recorded neural data while the participant was at rest with room noise minimized. These data were processed offline and used to assess signal quality. Neural signals were recorded using the NeuroPort data acquisition system (Blackrock Microsystems, Salt Lake City, UT) and were sampled at 30 kHz with a 750 Hz first order high pass filter [27]. Each time the recorded voltage signal crossed a pre-defined threshold of -4.5 times the baseline root-mean-square on each electrode, the time of the crossing and a 48-sample (1.6 ms) snippet of the signal, starting 11 samples before the crossing, was saved for offline analysis of signal quality.

### 2.5 Signal quality

Neural activity recorded at rest was processed offline in MATLAB (Natick, MA) to measure the number of high-amplitude recordings, peak-to-peak voltage, noise, and signal-to-noise ratio (SNR) over time. The peak-to-peak voltage was calculated individually for every snippet. Only the snippets with peak-to-peak voltages within the top 2% on a given electrode were used for analysis. This method was used to isolate the largest neural recordings and provide an unbiased estimate of the capacity of each electrode to record signals. More typical single-unit based analysis, especially over the long time periods of this study, could be subject to biases introduced by manual spike sorting [28]. We therefore selected analysis methods that did not require spike sorting. The largest 2% of the snippets for each electrode were averaged and the peak-to-peak voltage was calculated. If the firing rate of all the snippets on any electrode was less than 1.67 Hz or the peak-to-peak voltage of the averaged signal was less than 30 µV, the electrode was excluded from analysis. We used 1.67 Hz because it ensured we would have at least two isolated waveforms for analysis when using only the top 2% of snippets across a minute of recording. Electrodes containing snippets which had peak-to-peak voltages over 100 μV were considered electrodes that contained high-amplitude recordings. The noise metric was calculated as three times the standard deviation of the first five-time samples of all action potential snippets. As the first five values of each snippet capture time points prior to the threshold crossing event, they provided a metric for how much variance existed due to noise in the recordings. A similar method for calculating noise was used in a previous publication looking at motor unit stability [14]. SNR was calculated as the magnitude of the peak of the averaged waveform divided by the calculated noise.

The filter applied to the recorded neural data was changed at Day 200 from a fourth order 250 Hz high-pass Butterworth filter to a first order 750 Hz high-pass Butterworth filter to reduce the effects of stimulus artifact. The increased cut-off frequency resulted in an overall decrease in noise and, therefore, an increase in SNR [27]. Because of this, we separated data into pre-and post-filter change epochs for all signal quality analysis. All reported statistical results for signal quality are from data collected after the filter change.

### 2.6 Impedances

Electrode impedances at 1 kHz were measured at the beginning and end of each test session for every electrode. Impedances were measured using the impedance mode built into the NeuroPort patient cable data acquisition system (Blackrock Microsystems, Salt Lake City, UT), which involves delivering a 1 kHz, 10 nA peak-to-peak sinusoidal current for 1-s to each implanted electrode. For our analysis, we only used impedance recordings from the beginning of the sessions. This is because post-session impedances are affected by the application of ICMS, although these values return to normal the following test session, as has been noted previously [29].

### 2.7 Detection thresholds

Detection thresholds were calculated on at least two electrodes each day that stimulation was provided. One of these two was electrode 19 while the other electrode tested was taken sequentially from a list of seven electrodes (electrodes 8, 11, 28, 34, 40, 54, and 57). These electrodes were selected because they spanned both arrays and consistently generated perceptible sensations, but were otherwise arbitrarily chosen. Detection thresholds were calculated using a three-down, one-up staircase method [30], [31]. This involved a two-alternative forced choice task in which participant was presented with a stimulus pulse train during one of two 1-s intervals separated by a 1-s delay period and had to select which interval contained the pulse train. The amplitude of the stimulus was increased by 2 dB after an incorrect response and decreased by 2 dB after three correct responses. After five changes in direction, the trial was stopped and the threshold was calculated as the average of the last ten values before the end of the trial.

### 2.8 Data analysis and statistics

We tested for changes in signal quality, impedance, and detection thresholds over time using linear regression. Data points that had values more than three scaled median absolute deviations away from the median were excluded from analysis as outliers. Impedances and detection thresholds followed an exponential decay so the regressor, days post-implantation, was log-transformed prior to regression. Median data on stimulated electrodes in somatosensory cortex were compared to data on non-stimulated electrodes in motor cortex using ANCOVA. We used ANCOVA because it allowed us to assess significant differences between mean values on stimulated and non-stimulated electrodes by accounting for changes over time as a covariate, producing an ANCOVA adjusted means comparison. Additionally, we were able to assess the interaction of the two factors, time and stimulation condition, to determine if the changes over time were significantly different between stimulated and non-stimulated electrodes, producing an ANCOVA interaction term. The coefficients for slopes and interactions were considered to be significantly different than zero or from each other at the P < 0.05 level and highly significant at the P < 0.001 level. All p-values reported are for the coefficient of the regression slope unless otherwise indicated.

For detection thresholds measured across all electrodes at specific time points, we analysed the differences between blocks using Kruskal-Wallis tests and Tukey’s honestly significant difference (HSD) post-hoc tests. A non-parametric test was used because the detection data across electrodes were non-normal at each time point, as determined with an Anderson-Darling test. We quantified the spatial clustering of detection thresholds on the arrays using Moran’s I which is a method to assess spatial autocorrelation [32]. The weighting scheme considered only adjacent electrodes as neighbors. The significance for Moran’s I was determined using a z-test comparison to the predicted outcome of a random distribution. For visual clarity, interquartile ranges (IQRs) shown in figures were smoothed with a nine-point moving average filter with a triangular kernel. All statistical analysis was performed in MATLAB (Mathworks, Natick, MA).

## 3. Results

### 3.1. Recording stability

We quantified recording quality using four metrics: the number of high-amplitude recordings, peak-to-peak voltage, noise, and signal-to-noise ratio. Overall, we found that all signal quality metrics decreased significantly over time for both the stimulated (somatosensory) and non-stimulated (motor) electrodes. Importantly however, stimulated and non-stimulated electrodes were affected differently (Figure 1).

**Figure 1.**
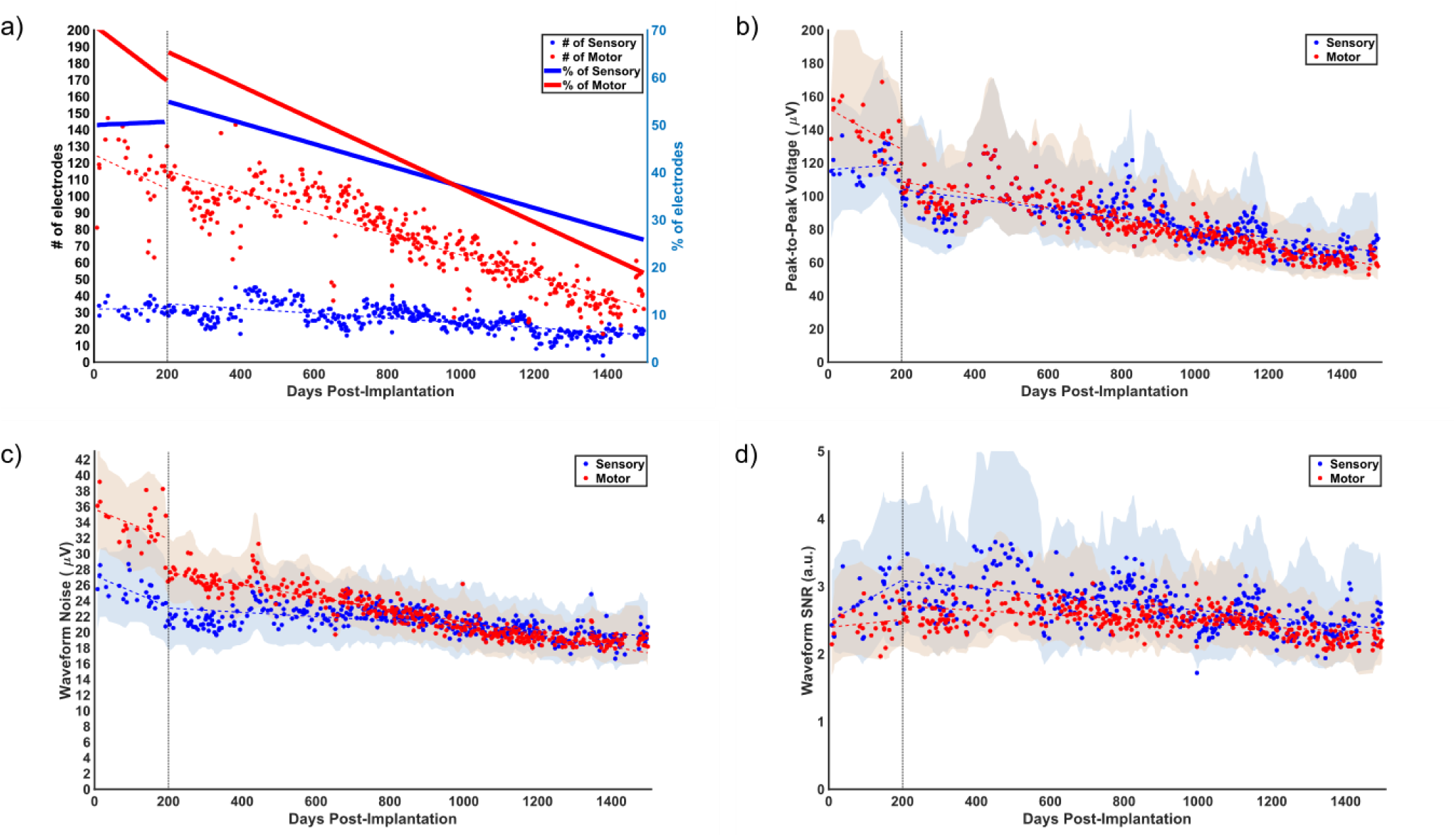
Signal quality over time. Blue points represent signal quality metrics from stimulated sensory electrodes and red points represent non-stimulated motor electrodes. Dotted lines represent regression before and after day 200. Shaded areas represent the smoothed interquartile range for each data set. The dotted vertical line marks day 200, when the filter order was changed. (**a**) The number of high-amplitude recordings since implantation. Each point represents the number of electrodes with high-amplitude recordings. The thick solid lines show the percentage of electrodes that had high-amplitude recordings for each group with a corresponding y-axis on the right. (**b**) Measured median peak-to-peak voltage since implantation in microvolts. (**c**) Measured median noise since implantation in microvolts. (**d**) Measured median signal-to-noise ratio since implantation.

The number of electrodes with high-amplitude recordings decreased significantly over time on both non-stimulated motor and stimulated sensory electrodes (P < 0.001, linear regression, Figure 1A). Interestingly, the stimulated electrodes with high-amplitude recordings decreased less over time (slope = -5 electrodes/year) than the non-stimulated electrodes with high amplitude recordings (slope = -23 electrodes/year). This difference was highly significant (P < 0.001, ANCOVA interaction) and reflects a 47% loss of high-amplitude signals on stimulated electrodes and a 72% loss of high-amplitude signals on non-stimulated electrodes from Day 200 to 1500. Early in the study, between Days 200 and 300, we recorded high-amplitude signals on 30±3 electrodes in somatosensory cortex and 101±10 electrodes in motor cortex. Late in the study, between Days 1400 and 1500, we recorded high-amplitude signals on 17±4 electrodes in somatosensory cortex and 38±9 electrodes in motor cortex. It should be noted that we record from 64 electrodes in somatosensory cortex and 176 electrodes in motor cortex, which favours recording a larger number of high-amplitude electrodes in the motor cortex. These results together demonstrate that stimulated electrodes better maintained high amplitude recordings over time than non-stimulated electrodes.

There were also highly significant differences in the changes in the peak-to-peak voltages, noise, and SNR on stimulated and non-stimulated electrodes (P < 0.001, ANCOVA interaction). Interestingly, the peak-to-peak voltage on stimulated electrodes decreased less over time (slope = -10.6 μV/year, P < 0.001, linear regression, Figure 1B) than on the non-stimulated electrodes (slope = -14.2 μV/year, P < 0.001, linear regression, Figure 1B) and the stimulated electrodes maintained a higher adjusted mean peak-to-peak voltage (P < 0.001, ANCOVA adjusted means). The noise on the stimulated electrodes also decreased less over time (slope = -1.0 μV/year, P < 0.001, Figure 1C) than on non-stimulated motor electrodes (slope = -2.9 μV/year, P < 0.001, Figure 1C) but the stimulated electrodes maintained a lower adjusted mean noise (P < 0.001, ANCOVA adjusted means).

This resulted in SNR decreasing more over time on stimulated electrodes (slope = -0.2 a.u./year, P < 0.001, Figure 1D) than on non-stimulated electrodes (slope = -0.1 a.u./year, P < 0.001, Figure 1D) due primarily to differences in how noise changed over time. This difference was highly significant (P < 0.001, ANCOVA interaction) and reflect a decrease in SNR of 23% on stimulated electrodes and 15% on non-stimulated electrodes from Day 200 to 1500. However, stimulated electrodes maintained a higher adjusted mean SNR than non-stimulated electrodes (P < 0.001, ANCOVA adjusted means). So, although the SNR decreased more over time on stimulated electrodes, a higher SNR was maintained throughout implantation than on non-stimulated electrodes. Early in the study, between Days 200 and 300, we recorded an SNR of 2.9±0.3 in somatosensory cortex and an SNR of 2.5±0.1 in motor cortex. Late in the study, between Days 1400 and 1500, the SNR had dropped by a small, but statistically significant amount to 2.4±0.5 in somatosensory cortex and 2.2±0.1 in motor cortex. This implies that significant differences in how SNR changed over time did not translate to practical differences in SNR, and in fact stimulated electrodes maintained higher SNR over time than non-stimulated electrodes.

To address the concern that charge itself might be associated with degradation in signal quality, we also investigated the signal quality metrics of a single electrode, electrode 19, which received the most stimulation of any electrode (Figure 2). The detection threshold for this electrode was tested during every stimulation session and this electrode was additionally the most commonly used electrode for bidirectional BCI tasks. Although the intersession variance was much larger for this single electrode than the median data, neither the peak-to-peak amplitude (P = 0.92, ANCOVA interaction, Figure 2A) nor SNR (P = 0.22, ANCOVA interaction, Figure 2C) were significantly different over time compared to all stimulated electrodes. However, the baseline noise decreased more on electrode 19 than the rest of the stimulated electrodes (P < 0.001, ANCOVA interaction, Figure 2B). This resulted in an adjusted mean noise that was significantly lower and an adjusted mean SNR that was significantly higher (P < 0.05, ANCOVA adjusted means) than the adjusted mean of the median noise and SNR for all stimulated electrodes. Next, we compared electrode 19 to the non-stimulated electrodes in motor cortex and found that the magnitude of the decrease in noise on electrode 19 over time (the slope) was not significantly different than the decrease in the median noise on the non-stimulated electrodes over time (P = 0.7786, ANCOVA interaction). However, the noise itself on electrode 19 was significantly lower than the median noise on non-stimulated electrodes (P<0.001, ANCOVA adjusted means). Overall, the noise on stimulated electrodes decreased more quickly than the noise on non-stimulated electrodes. That this effect did not appear to be driven by the amount of stimulation (electrode 19 was no different than non-stimulated electrodes), suggests that other factors such as electrode material and impedance could have been the cause.

**Figure 2.**
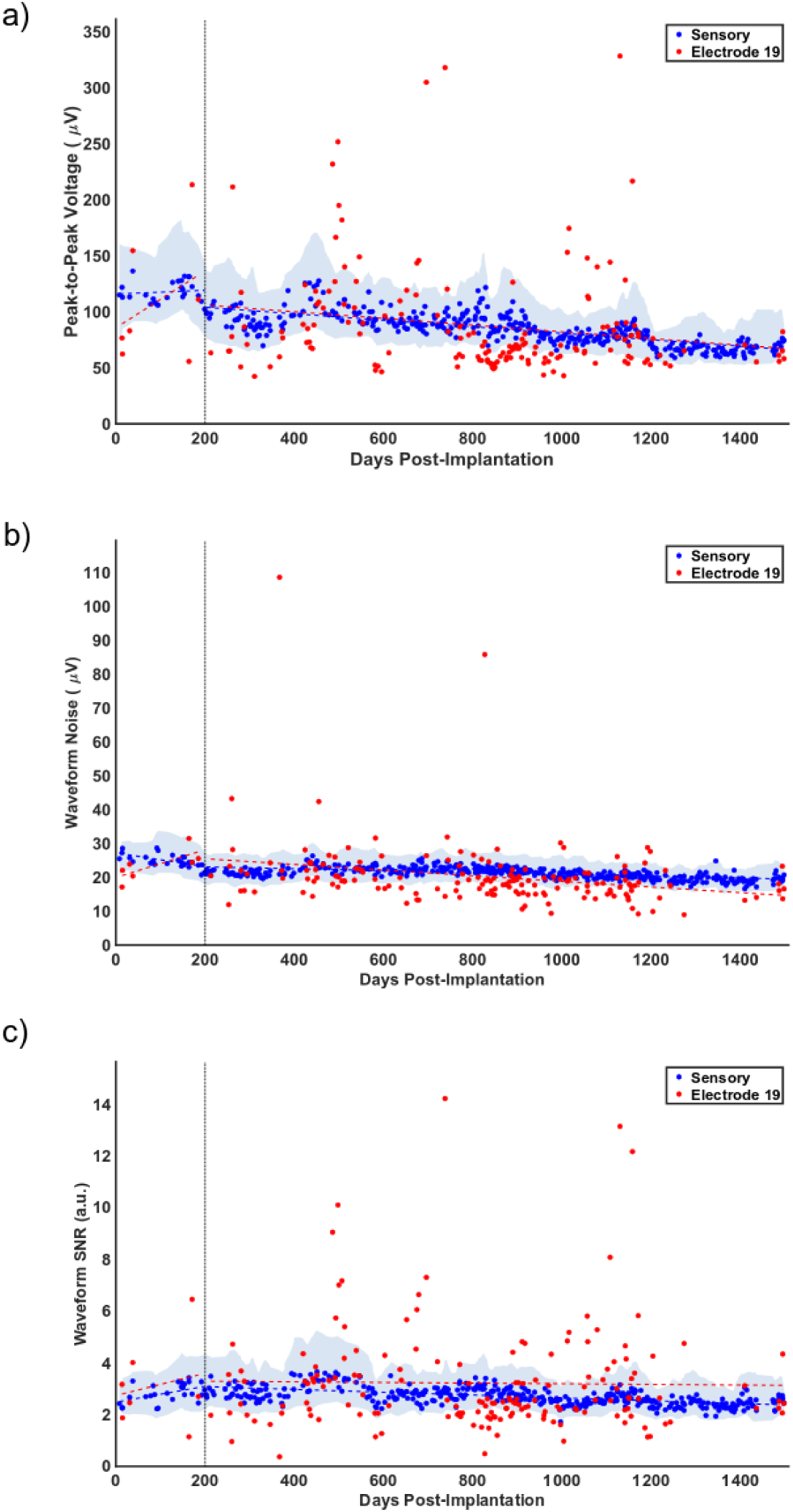
Signal quality plots on a single electrode that received the most stimulation. Points represent median values from test days. Dotted lines show regression across all data. The dotted vertical line marks day 200, when the filter order was changed. (**a**) The measured peak-to-peak voltage since implantation in microvolts. (**b**) Measured noise since implantation in microvolts. (**c**) Measured signal-to-noise ratio since implantation.

### 3.2 Impedance Stability

There was a large increase in impedances at 1 kHz following implantation on both motor electrodes (median pre-implant impedance: 447.5 kOhms; median Day 7 impedances: 1396 kOhms) and sensory electrodes (median pre-implant impedance: 74.5 kOhms; median Day 7 impedances: 533 kOhms). We assessed whether there was any significant change in the electrode impedances following implantation over 1500 days using linear regression (Figure 3A). We found that there was a highly significant decrease in electrode impedance over time on both the stimulated and non-stimulated electrodes (P < 0.001). Additionally, there was a highly significant difference in the changes in impedance between stimulated and non-stimulated electrodes (P < 0.001, ANCOVA interaction) with a greater decrease in impedance over time on the non-stimulated electrodes. Over 1500 days, the impedance on the stimulated electrodes decreased by 79% while the non-stimulated electrode impedances decreased by 85%. It should be noted that the median impedances on the platinum electrodes in motor cortex were initially much higher than the SIROF electrodes in somatosensory cortex [33].

**Figure 3.**
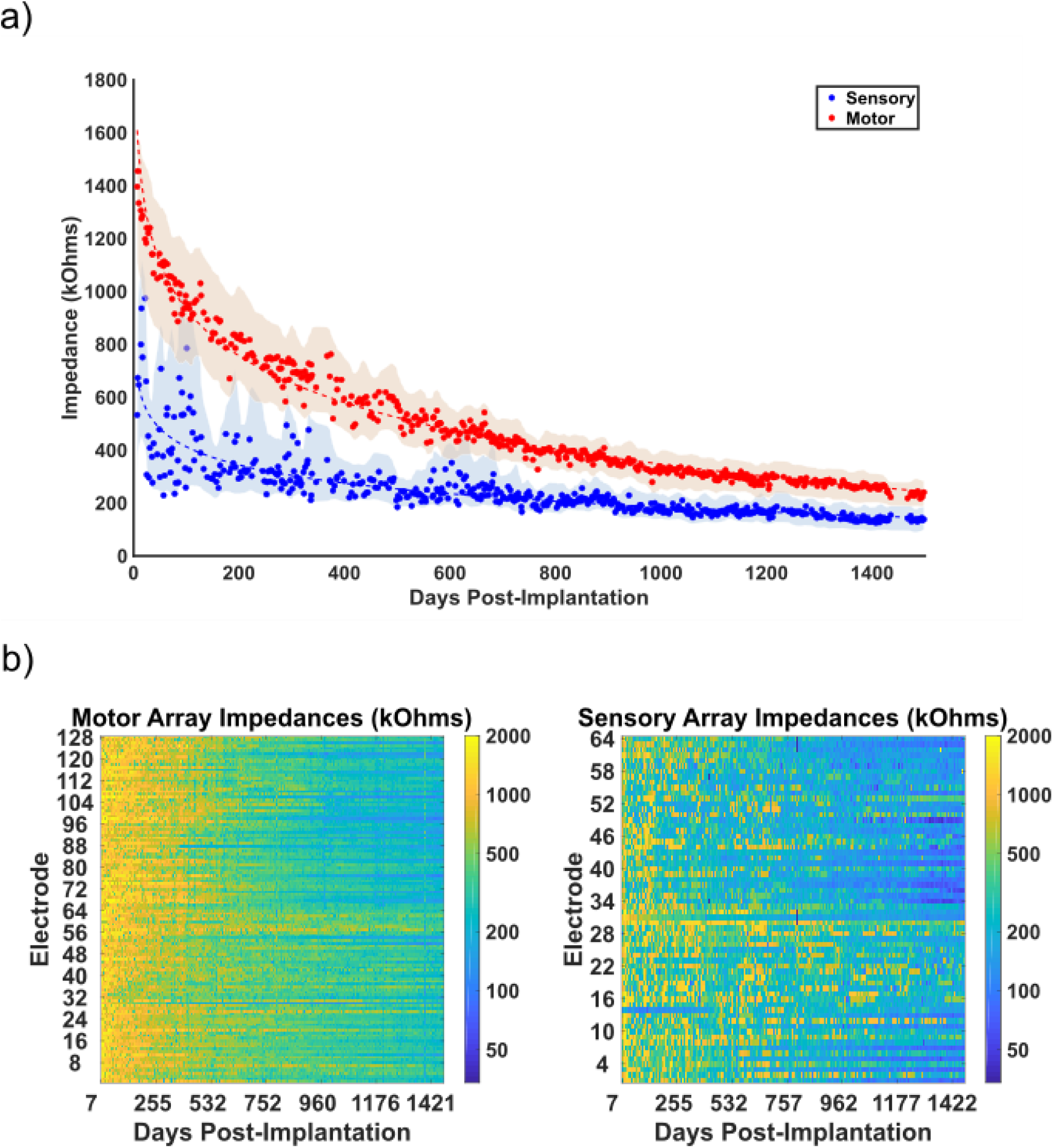
Electrode impedance over time. (**a**) Plots for impedances of all electrodes plotted against days since implantation for the stimulated sensory array (blue) and the non-stimulated motor array (red). Each point represents the median impedance for the test day. Dotted lines represent the linear regression of the data with log-transformed days. Shaded areas represent the smoothed interquartile range for each data set. (**b**) Heatmaps for impedances of all electrodes plotted against days since implantation for the motor array (left) and sensory array (right). Color maps were log transformed to better illustrate changes over time.

Impedance measurements on individual electrodes were often highly variable from day to day, making the comparison of individual electrodes to group median values impossible. Electrode 19, the most stimulated electrode, was one electrode with high intersession variance, making comparisons of its impedance to other stimulated or non-stimulated electrodes uninformative. We suspect that this was due to issues related to the specific connector and headstage used to measure impedances.

### 3.3 Detection Threshold Stability

Detection thresholds were measured in four different time ranges: near Day 100, Day 500, Day 1000, and Day 1500. Due to limits on the participant’s time in the lab for each test session, these measurements were typically taken across multiple test sessions. Nearly all of the implanted electrodes (n=64) were tested within each time range with 54 electrodes tested around Day 100, 61 electrodes around Day 500, 58 electrodes around Day 1000, and 53 electrodes around Day 1500. Some electrodes were excluded from detection due to high voltages during the interphase period at the beginning of the test session.

Surprisingly, we saw that over the course of the study, the median detection thresholds decreased by about two-thirds, from 31.5 μA at Day 100 (IQR: 22.8-49.6 μA), to 15.8 μA at Day 500 (IQR: 9.0-24.9 μA), 12.4 μA at Day 1000 (IQR: 8.1-18.0 μA), and 10.4 μA at Day 1500 (IQR: 7.4-16.6 μA) (Figure 4A). We found that detection thresholds around Day 100 were significantly higher than Days 500, 1000, and 1500 (P < 0.001, Kruskal-Wallis test with Tukey’s HSD post-hoc test) but other sessions were not significantly different from each other (P > 0.05, Kruskal-Wallis Test with Tukey’s HSD post-hoc test). We also assessed whether the detection thresholds were spatially organized on the arrays (Figure 4B) and found that for all test ranges except Day 100, there was significant clustering (P < 0.05, Moran’s I with z-test). This indicates that for most of the implant duration, the detection thresholds were not randomly distributed across the array and electrodes with high thresholds tended to be near other electrodes with high thresholds.

**Figure 4.**
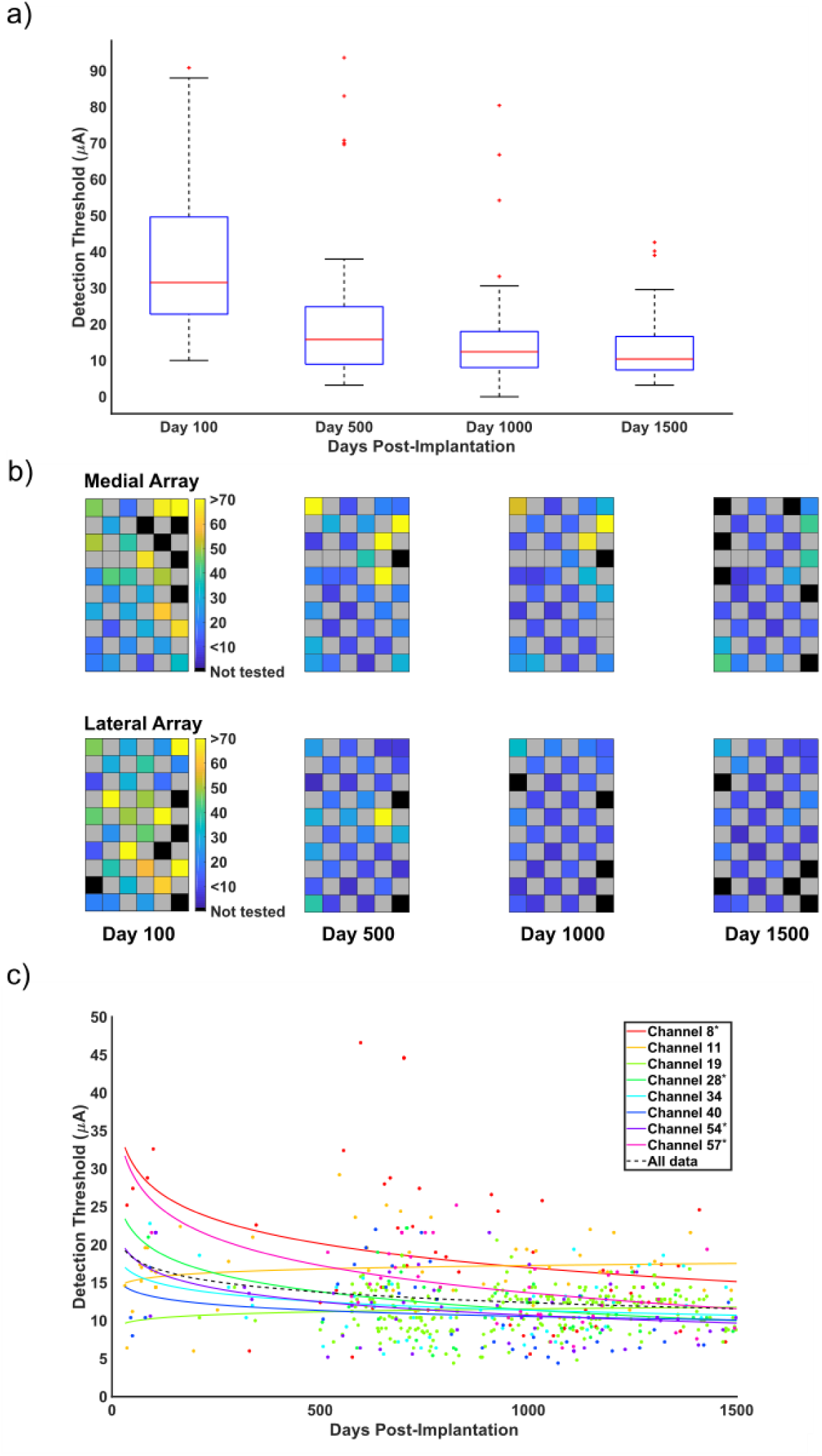
Detection thresholds for stimulated electrodes since implantation. (**a**) Box plots for the median threshold values from each test range. The four time ranges are shown chronologically from left to right. (**b**) The measured detection thresholds for each electrode plotted to the array as a heat map. Black spaces represent electrodes that were not tested. Grey spaces represent electrodes that were not wired due to technical constraints. The color bars show the detection threshold values in microamperes. (**c**) The measured detection thresholds for eight electrodes are plotted over time. Each point represents an individual threshold for a single electrode. The color of the point indicates a specific electrode as indicated in the legend. Asterisks on the legend indicate electrodes with statistically significant changes over time. Lines were fit to the data for each electrode using regression of the log-transformed data. The fits were reverse transformed for plotting. The dotted black line shows the regression for the aggregate detection data.

Finally, we measured the detection thresholds on a subset of electrodes more frequently throughout the study. These electrodes spanned the two sensory arrays but were otherwise chosen arbitrarily. Changes over time were quantified with regression of the individual electrode detection data using a log-transformed time axis. Of the eight tested electrodes, the detection threshold on four electrodes decreased significantly over time (P < 0.05, Figure 4C, indicated with *) while no electrode had significant increases. The analysis of the group data in Figure 4A showed that after Day 500 there was no further decreases in detection thresholds. To examine whether this was true on the eight electrodes where we had considerably more measurements, we performed linear regression for data collected after Day 500. We found that on five electrodes there was a significant change in detection thresholds after Day 500 (P < 0.05). Four of these were significant decreases in thresholds and one was a significant increase. However, these changes were all small, with slopes of -5.4, -4.2, -3.2, -2.2, and 2.0 μA/year.

## 4. Discussion

Microstimulation in the somatosensory cortex of a person using SIROF-coated Utah microelectrode arrays can evoke detectable sensations for 1500 days. We found that stimulated electrodes maintained a higher SNR than non-stimulated electrodes throughout the length of the implant, although the SNR on stimulated electrodes declined at a faster rate. In fact, stimulated electrodes were far more likely to retain the capacity to record high-amplitude neurons after 1500 days than electrodes that had never been stimulated. Indeed, 50% of the stimulation electrodes recorded high-amplitude neurons within the first 100 days and 25% recorded high-amplitude neurons in the last 100 days, representing a 50% loss. However, on non-stimulated electrodes in the motor cortex, 70% recorded high-amplitude neurons within the first 100 days, but just 20% recorded high-amplitude neurons in the last 100 days, representing a 70% loss. Electrode impedances for stimulated electrodes had a significantly smaller decrease over time. Finally, and perhaps most importantly, stimulation detection thresholds continually decreased for 1500 days, indicating an improvement in the ability to elicit sensations with ICMS.

### 4.1 Stimulation safety limits

The safety limits on the stimulus parameters used in this study, such as pulse amplitude and frequency were based on a series of studies in non-human primates investigating the effects of stimulation on safety at the electrode tissue interface [20], [21]. However, these pulse parameters have not always been shown to be safe [18], [19], [34]–[40]. Of particular note, a study of chronic microstimulation in the cat cortex by McCreery et al. showed that continuous stimulation at even at the low level of 4 nC/phase (equivalent to a stimulation amplitude of 20 µA with the 200 μs pulse width we used in this study), led to a loss of neurons around the electrode tips [39]. While we regularly exceeded this stimulation amplitude and use the full amplitude range up to 100 µA on implanted electrodes, we limited the duty cycle to a maximum of 15 seconds of continuous stimulation, at which point stimulation would be disabled for an equivalent amount of time. This may be why stimulation with these parameters did not lead to measurable deleterious effects on electrodes or evoked sensations.

### 4.2 Recording signal quality

It is well known that signal quality deteriorates on recording electrodes over long periods of time [5], [10]–[12], [14], [15]. In this study, we found similar decreases in signal quality over time on both stimulated and non-stimulated electrodes. Importantly, across 1500 days we saw a 37% decrease in the peak-to-peak voltage on the stimulated electrodes, while the peak-to-peak voltage on non-stimulated electrodes decreased by 46%. For comparison, Chestek et al. 2011 reported a 28.2% and 47% decrease in single-unit amplitude in two implanted non-human primates within the first two months after implant while Barrese et al. 2013 showed an average decrease in the signal amplitude of about 12% across 27 implanted non-human primates over 1000 days on viable electrodes. However, about half of electrodes were excluded from analysis because they were considered not viable, meaning the peak-to-peak voltage fell below 40 μV. Our results are consistent with these and other studies showing that signal quality decreases over time. However, interestingly we saw a significantly greater decrease in both the peak-to-peak voltage and the number of high-amplitude recordings on non-stimulated electrodes. These findings suggest that, at a minimum, ICMS did not additionally contribute to decreases in recording quality over time and may actually improve the longevity of high-amplitude recordings.

Previous work has posited that stimulation can rejuvenate the electrode-tissue interface, resulting in increased SNR through an improvement in signal amplitude and decreased impedances [41], [42]. This rejuvenation effect could be related to the preservation of high amplitude recordings on stimulated electrodes that we observed. However, our results showed a steeper decline in SNR over time on stimulated electrodes compared to non-stimulated electrodes, although the SNR itself was higher on stimulated electrodes than non-stimulated electrodes. Furthermore, we saw that the impedance on stimulated electrodes declined at a slower rate than the impedances on non-stimulated electrodes. Impedances however were difficult to compare between stimulated and non-stimulated electrodes since these electrodes contained different materials with different impedance properties, and stimulated electrodes maintained lower impedance values throughout. The previous work suggesting a rejuvenation effect was focused on the short-term effects of stimulation on signal properties, with a maximum tested duration of eight days. Changes in the signal and impedance over this time scale may not reflect expected changes over many years. Another substantial difference between the previous work and this current study is the stimulus protocol. In the previous work, stimulation was held constant at 1.5 V for four seconds, and this pulse was only delivered once prior to all signal and impedance measurements. Here, our stimulus parameters consisted of more typical stimulation protocols, including charge-balanced biphasic pulses with an initial cathodic phase lasting 200 μs. Stimulus trains were delivered at a maximum of 300 Hz for up to 15 seconds at a maximum amplitude of 100 μA over many years. The substantial differences in our stimulus protocols makes it difficult to assess similarities or differences in the results.

### 4.3 Impedances

Impedances on electrodes have been shown to increase immediately after implantation but decrease over long periods of time [10], [34]–[36]. Our results similarly showed an increase from pre-implant impedance values and then a decrease in impedances across the 1500 days of implantation. While the impedances of the non-stimulated electrodes implanted in the motor cortex had a significantly greater decrease than the stimulated electrodes in the somatosensory cortex, this effect is likely related to the difference material properties of the electrodes in these two locations. Sputtered iridium-oxide electrodes have significantly lower impedances than platinum electrodes as a result of their significantly larger surface area, due in large part to the sputtering deposition process [33]. As a result, the impedances on the stimulation electrodes were much lower than the platinum electrodes shortly after implantation and therefore did not have as much range to decrease. Nevertheless, these data demonstrate that over 1500 days, including nearly 378 days of ICMS, the electrode impedances were well within ranges suitable for microstimulation. Increasing impedances, potentially as a result of electrode damage resulting from stimulation itself, would result in higher voltages at the electrode-tissue interface which could damage tissue [26], or exceed the compliance voltage limits of the stimulator itself. Although differences in the changes over time were significant, the difference in the percent change between stimulated and non-stimulated electrodes was small and may not be practically meaningful.

### 4.4 Detection thresholds

Perhaps the most supportive demonstration of the long-term utility of microstimulation in somatosensory cortex was the observation that that sensory percepts could be consistently evoked over the length of implantation, with detection thresholds decreasing over time. Non-human primate studies had demonstrated that sensations could be evoked over periods of months without behavioral deficits using stimulation parameters higher than those used in this experiment [20], [34]–[36], [40], [43]. This is, however, the first time a human participant has been implanted and tested with this protocol and it is promising for bidirectional BCIs that percepts can be reliably evoked and that detection thresholds themselves decrease over long periods time. Detection of stimulation could improve over time for a variety of reasons. As we mentioned previously, stimulation could possibly provide rejuvenating effects on neural tissue, which could result in more responsive neurons. Stimulation over many years could also possibly result in neural plasticity resulting in stronger sensitivity to stimulation [44]. Additionally, the participant’s familiarity with the stimulus detection task could have resulted in increased behavioural sensitivity. We also demonstrated that the detection thresholds were spatially clustered on the electrode arrays. There are several possible explanations for this effect. First, if the arrays were not perfectly perpendicular to the cortical surface, or if the cortical surface was curved under the electrode, individual electrode tips on different parts of the array could be at different depths in the cortex, a factor which is known to have an effect on detection thresholds [45], [46]. Additionally, localized tissue reactions could potentially affect the microenvironment around individual electrodes [47] and adjacent electrodes or different locations in somatosensory cortex could be differentially responsive to stimulation. Further work is needed to explore the mechanisms of increased stimulus sensitivity as well as spatial clustering of thresholds.

### 4.5 Study limitations

Although these results are supportive of the long-term utility of microstimulation in the somatosensory cortex as a component of implanted BCIs, we have only tested this in one participant. Future implantations will need to be similarly evaluated to determine if signal quality and detection thresholds are stable over long periods of time. Furthermore, this study gives a general overview of stability on electrodes that have been stimulated as compared to electrodes which have not been stimulated but does not delve deeply into how the amount of stimulation delivered on each electrode might impact signal quality, impedances, or detection thresholds. We have provided results for a single electrode which was stimulated much more frequently than any other electrode due to its involvement in many of our bidirectional BCI paradigms. Its signal quality over time did not deviate significantly from other stimulated electrodes except for noise, which had greater decreases over time. Another possible limitation is that the stimulation delivered as part of this experiment may be much less than what would be provided in a deployable BCI system. However, as stated previously, we have not seen any notable differences between electrodes based on the amount of stimulation they have received. As mentioned previously, the electrodes that were provided stimulation were coated with iridium oxide while the electrodes that were not provided stimulation were coated with platinum. If there are differences in how these materials degrade over time regardless of stimulation, comparisons can be difficult to make. For better comparison, future implants should have iridium oxide coated tips on all electrodes. Another consideration is that stimulated electrodes were implanted in somatosensory cortex while non-stimulated electrodes were implanted in motor cortex. Neurophysiological differences in these two areas could possibly affect the results. For example, layer 5 of motor cortex contains large Betz cells while layer 4 of somatosensory cortex contains smaller pyramidal cells [25]. This difference in cell type is the likely cause of the initially high-amplitude signals recorded from motor cortex. Whether neuroanatomical differences in motor and somatosensory cortex could have differential effects on the response to microstimulation or the presence of implanted devices over many years is unknown.

### 4.6 Conclusion

Overall, these findings indicate that electrodes receiving intracortical microstimulation in somatosensory cortex can maintain recording quality as well, if not better, than non-stimulated electrodes in motor cortex and continually elicit sensory percepts with stimulation over 1500 days. While this time frame may still be a relatively short in the context of a life-long implantation, demonstrating stability at these time points is a necessary step to support the continued development of these systems. These findings are promising for the field of neural prostheses and specifically sensory restoration and indicate that implanted microelectrodes for sensory restoration is plausible for clinical application.

## Data Availability

Data supporting these findings as well as software routines to analyze these data are available from the corresponding author upon reasonable request.

## Acknowledgements

We would like to acknowledge Nathan Copeland for his extraordinary commitment and effort in relation to this study and insightful discussions with the study team; Debbie Harrington (Physical Medicine and Rehabilitation) for regulatory management of the study; Angelica Herrera for her help with data acquisition. This work was supported by the Defense Advanced Research Projects Agency (DARPA) and Space and Naval Warfare Systems Center Pacific (SSC Pacific) under Contract N66001-16-C-4051 and the National Institute Of Neurological Disorders And Stroke of the National Institutes of Health under Award Numbers UH3NS107714 and U01NS108922. Any opinions, findings and conclusions or recommendations expressed in this material are those of the authors and do not necessarily reflect the views of DARPA, SSC Pacific, or the National Institutes of Health.

